# How has the municipal availability of the *GeneXpert*^®^*MTB/RIF* system affected the detection of drug-resistant tuberculosis in Brazil?

**DOI:** 10.1101/2023.03.07.23286916

**Authors:** Jhancy Rocío Aguilar-Jiménez, Daniele Maria Pelissari, Fredi Alexander Diaz-Quijano

## Abstract

**Objective:** To evaluate the association between the availability of *GeneXpert*^*®*^*MTB/RIF* in municipalities and the proportion of people who have access to this diagnostic technology for tuberculosis (TB), as well as the resistance detected by the surveillance system in Brazil.

**Methods:** We analyzed 4,998 Brazilian municipalities that reported 432,937 new TB cases between 2015 and 2020. We compared municipalities with and without the availability of *GeneXpert*^*®*^*MTB/RIF* regarding the prevalence of effective access to *GeneXpert*^*®*^*MTB/RIF* diagnosis and that of detected resistance.

**Results:** Municipalities with at least one *GeneXpert*^*®*^*MTB/RIF* system had three times (95%CI 2.9-3.0) the access to diagnostic tests and 80.4% (95%CI 70.6%-90.2%) higher detection of resistance, compared to municipalities without this technology. We estimate that there have been 2,110 cases of undetected resistance during this period in the country.

**Conclusions:** The availability of *GeneXpert*^*®*^*MTB/RIF* system in the municipality increased the sensitivity of the surveillance for detecting TB resistance.

**Public Health implications:** It is a priority to strengthen laboratory networks and narrow the gap in access to rapid diagnosis in remote areas to improve the detection and control of drug-resistant tuberculosis.

## INTRODUCTION

Drug-resistance is one of the main issues in eradicating tuberculosis (TB) as a public health problem ^1,2^. Moreover, accelerating diagnoses and initiating specific treatments represent a challenge to public health systems ^2,3^. The *GeneXpert*^*®*^*MTB/RIF* test allows the detection of both *Mycobacterium tuberculosis* genetic materials and mutations that cause resistance to rifampicin ^1,4^. This rapid test has been progressively introduced in Brazil, but is not yet available to all municipalities. The Brazilian diagnostic network is universally accessible and equitable, therefore, the municipalities served by the *GeneXpert*^*®*^*MTB/RIF* system are expected to support diagnosis to patients from locations that do not have it. However, aspects such as transporting samples from remote areas could represent a geographical barrier to this access ^5,6^. Thus, we evaluated the extent to which the availability of *GeneXpert*^*®*^*MTB/RIF* could influence access to this diagnostic technology and the detection of drug-resistance in new cases of TB in Brazil.

## METHODS

A total of 4,998 out of 5,570 Brazilian municipalities reported new TB cases between January 2015 and December 2020. Our data was updated up to August 2021, therefore, each of the cases had a follow-up of at least seven months until the final classification of resistance. Information from 2014 (or earlier) was not taken into account because it was the year of implementation of the *GeneXpert*^*®*^*MTB/RIF* in Brazil. Reported TB cases were retrieved from the Information System for Notifiable Diseases (SINAN), population estimates by municipality from the Brazilian Institute of Geography and Statistics (IBGE), and the installation dates of the *GeneXpert*^*®*^*MTB/RIF* system were obtained from the Ministry of Health of Brazil. For each municipality, the proportion of tuberculosis cases with access to *GeneXpert®MTB/RIF* and those with resistance detected by any method (either by *GeneXpert®MTB/RIF* or by sensitivity testing) was calculated. We estimated the prevalence ratios between municipalities with and without *GeneXpert*^*®*^*MTB/RIF*, with 95% confidence intervals, for the proportion of effective access to GeneXpert®MTB/RIF and the proportion of resistance detected.

We explored the number of devices per capita (100,000 inhabitants) and by TB cases (1,000 cases), considering the time of device use in the municipality since its installation and approximating, from tercile cutoffs to whole numbers. After, based on the pseudo R2 of the Poisson regression, we identified that the ratio of device to TB cases predicted the proportion of detected resistance better than the ratio of device to population. Therefore, we quantified the relative under-detection of estimated resistance considering as a reference the prevalence detected in the municipality group with the highest number of devices per case. Analyses were performed using Stata version 16.0 (StataCorp LP, College Station, TX) and Epidat 4.2 ^®^, the maps were designed in a QGis^®^ software.

Interpretation: The prevalence of resistance detected (*Rd*) is equal to the product of the prevalence of true resistance (*Rv*) multiplied by the sensitivity (*S*) of the system for detection. When two groups are compared, we calculate the prevalence ratio (PR) of resistance detected, as follows:

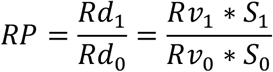

Where subscript 1 to the values of the interest group and subscript 0 to the reference group. Assuming that resistance is relatively constant across groups of municipalities (*Rv*_*0*_ ≈ *Rv*_*1*_), the prevalence ratio (*RP*) of resistance of patients identified in municipalities where *GeneXpert*^*®*^*MTB/RIF* was available compared to those who do not have it, we interpret as a ratio of sensitivities for detection of resistance in TB (*RP*≈ *S*_1_*/S*_0_).

### Ethics statement

Data used in this study do not include the personal identification of TB patients. All data analyzed are publicly available in Brazil. According to Resolution No. 510 of the Brazilian National Health Council, research conducted exclusively with public data is not evaluated by an institutional review board.

## RESULTS

A total of 270,069 cases of TB were reported among the 136 municipalities equipped with the *GeneXpert*^*®*^*MTB/RIF* technology. Out of these cases, 41% (n=110,615) were diagnosed by *GeneXpert*^*®*^*MTB/RIF* method, and resistance was reported in 4,916 cases (1.8%). In municipalities deprived of a *GeneXpert*^*®*^*MTB/RIF* system, (n=4,862), 162,868 cases of TB were reported, of which 13.8% (n=22,461) had access to diagnosis with *GeneXpert*^*®*^*MTB/RIF*, and resistance was detected only in 1.0% (n=1,643) of cases. Thus, compared with municipalities without *GeneXpert*^*®*^*MTB/RIF*, those with at least one device exhibited 3.0 times (95% CI 2.9-3.0; p<0.001) access to that diagnostic technology, and 80.4% higher (95% CI 70.6%-90.2%; p<0.001) resistance detection.

In the subgroup of municipalities with at least two devices per 1000 cases of TB, the prevalence of detected resistance was 2.0%, which corresponded to almost double (PR 1.99 95% CI 1.87%-2.10%) of those registered in the municipalities without a *GeneXpert*^*®*^*MTB/RIF* (1.01%) system. Extrapolating the proportion of resistance observed in the municipalities with greater availability of the *GeneXpert*^*®*^*MTB/RIF* technology, we estimated that there was a total of 2.110 cases of non-detected resistance during the period, which corresponded to a relative underreported resistance of 24.3% (95% CI 21.98%-26.71%). The underreporting would be at 49.6% in municipalities without *GeneXpert*^*®*^*MTB/RIF*, 27.2% in those with less than 1 device/1000 cases, and 20.8% in municipalities with 1 to less than 2 devices/1000 cases (Table 1).

**Table 1.** Prevalence of access to *GeneXpert MTB/RIF* system and resistance detected by the surveillance system in Brazilian municipalities classified according to the availability of devices, 2015-2020.

| Number of<br>*GeneXpert*<br>MTB/RIF<br>devices /1000<br>TB cases | Number of<br>municipalities | Total<br>TB<br>cases | Number of<br>Cases with<br>*GeneXpert*<sup>®</sup><br>MTB/RIF (%) | Number of<br>detected<br>resistant<br>cases<br>(%) | Relative<br>underreporting of<br>resistance -%<br>(95% CI) | Potentially non-detected resistant<br>cases (95% CI) |
| --- | --- | --- | --- | --- | --- | --- |
| 0 | 4,862 | 162,868 | 22,461 (13.8) | 1,643 (1.01) | 49.6 (46.6-52.5) | 1,618 (1,519-1,711) |
| > 0 to < 1 | 40 | 63,075 | 22,483 (35.6) | 920 (1.46) | 27.2 (21.7-32.2) | 343 (274-407) |
| ≥ 1 to < 2 | 44 | 35,860 | 14,987 (41.8) | 569 (1.59) | 20.8 (13.5-27.4) | 149 (97-197) |
| ≥ 2 | 52 | 171,134 | 73,145 (42.7) | 3,427 (2.0) | - | Reference |
| **Total** | 4,998 | 432,937 | 133,076 (30.7) | 6,559 (1.52) | 24.3 (21.9-26.7) | 2,110 (1,905-2,315) |

Regarding the studied geographic distribution, we observed a trendy pattern between the proportions of effective access to *GeneXpert*^*®*^ *MTB/RIF* (Figure 1a) and the proportion of detected resistance (Figure 1b) in municipalities provided with *GeneXpert*^*®*^*MTB/RIF* system or those within their geographical coverage.

**Figure 1a.**
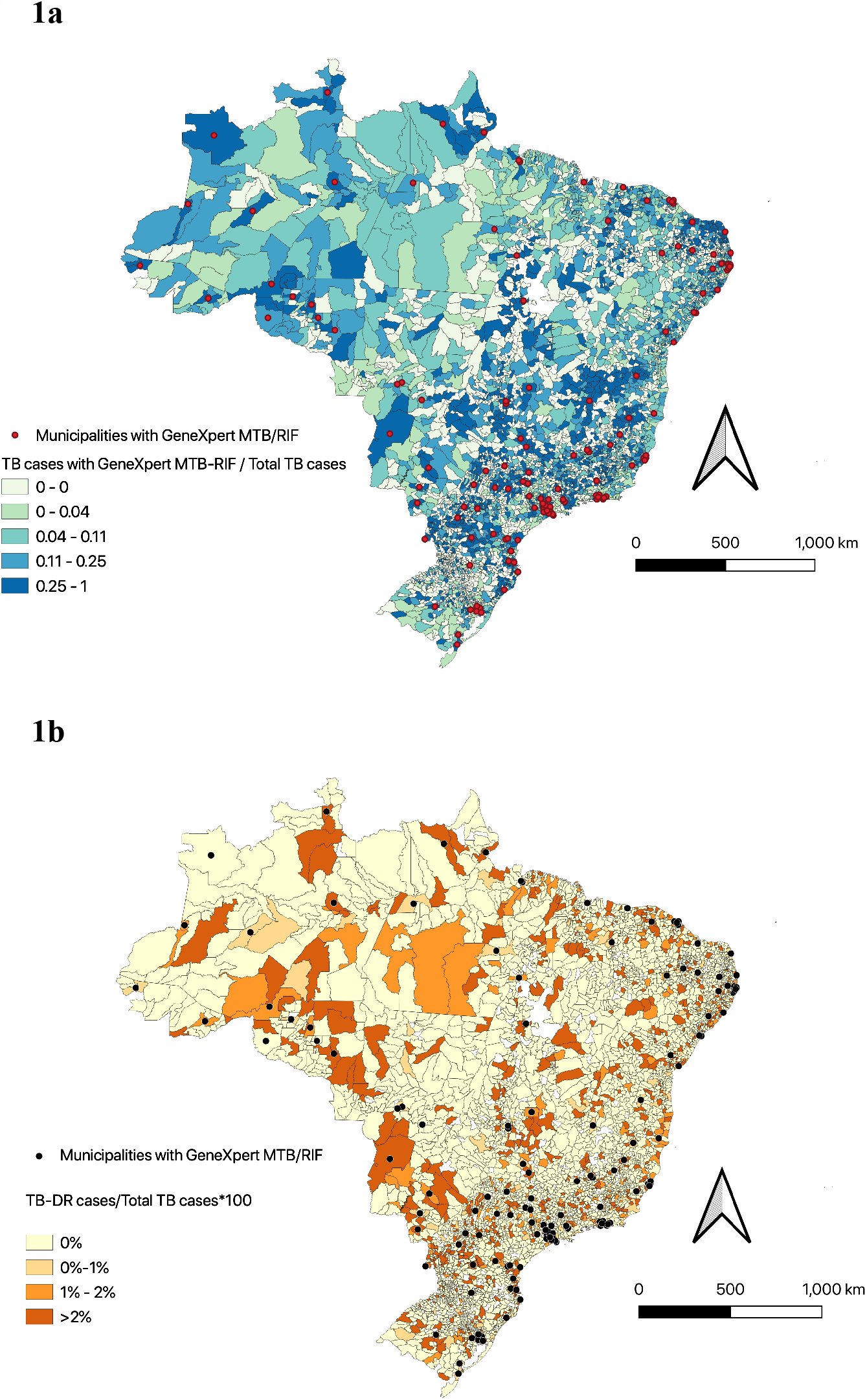
Distribution of the proportion of TB cases with access to *GeneXpert*^*®*^MTB/RIF. Figure 1b. Distribution of the proportion of drug-resistant TB cases.

## DISCUSSION

Through this national study, we described the variability of access to advanced TB diagnostic technologies and estimated the probable increase in sensitivity of resistance detection associated with its availability among municipalities.

Previous studies reported that implementation of *GeneXpert*^*®*^*MTB/RIF* reduced time to treatment initiation and increased detection of drug-resistant TB ^7,8^. Our results suggest that municipalities with this technology had a higher sensitivity to detect resistance. In addition, we observed that an optimal sensitivity would require at least one device for every 500 cases of tuberculosis.

We interpret the comparison of detected resistances as reflecting differences in sensitivity. However, this implies assuming a constant resistance prevalence across municipalities, which is a strong assumption because there are different determinants that could also affect the resistance frequency ^9^. Therefore, it would be advisable for future studies to consider individual and contextual determinants when evaluating the introduction of diagnostic technologies for detection resistance.

Previous studies suggested that the sensitivity to drug-resistant TB identification is only 46.6% in Brazil ^10^. The present study did not intend to calculate the sensitivity, but to quantify the non-detected resistance cases due to a lack of proper technology. However, we believe that even the municipalities with the highest availability of *GeneXpert*^*®*^*MTB/RIF* system may have some underreported cases.

#### Public health implications

Strengthening diagnostic networks by expanding *GeneXpert*^*®*^*MTB/RIF* availability, therefore reducing the gap in access to rapid diagnosis in remote geographic areas should be deemed necessary. These findings may align with global recommendations for increased funding to achieve universal access ^11,12^. They also contribute to the approach proposed by the Brazilian Ministry of Health to improve indicators, identifying areas for restructuring the expansion of laboratory and diagnostic networks, keeping as a target the eradication of TB in Brazil by 2035 ^10,12^.

## CONCLUSIONS

This study suggested that the availability of *GeneXpert*^*®*^*MTB/RIF* system within municipalities increases the sensitivity for the detection of drug-resistant TB. Thus, we expect these results to be taken into account for the adjustment of resistance indicators and to planning the expansion of the use of *GeneXpert*^*®*^*MTB/RIF* system, alongside cost-effectiveness studies.

## Data Availability

All data used in this study are openly accessible and available through the sources listed in the manuscript.

https://portalsinan.saude.gov.br/tuberculose

## Conflict of interest and human participant protection statement

We declare that we have no potential conflict of interest. Data used in this study do not include the personal identification of TB patients. All data analyzed are publicly available in Brazil.

## Acknowledgments

We thank Patricia Marques Moralejo Bermudi for her support in the creation of the maps in QGis^®^. This work had no specific funding. However, FADQ is beneficiary of a fellowship for research productivity from the National Council for Scientific and Technological Development - CNPq, process/contract identification: 312656/2019-0.

